# Artificial Intelligence-Guided Molecular Determinants of PI3K Pathway Alterations in Early-Onset Colorectal Cancer Among High-Risk Groups Receiving FOLFOX

**DOI:** 10.1101/2025.08.18.25333929

**Authors:** Fernando C. Diaz, Brigette Waldrup, Francisco G. Carranza, Sophia Manjarrez, Enrique Velazquez-Villarreal

## Abstract

**Background:** Early-onset colorectal cancer (EOCRC), defined as diagnosis before age 50, is rising rapidly and disproportionately affects high-risk populations, particularly Hispanic/Latino (H/L) individuals, who experience the steepest increases in incidence and mortality. While prevention and screening strategies have curbed late-onset CRC rates, EOCRC remains outside standard screening guidelines and is projected to become the leading cause of cancer-related death in individuals aged 20–49 by 2030. FOLFOX (folinic acid, fluorouracil, and oxaliplatin) is a standard first-line therapy for microsatellite stable (MSS) CRC lacking actionable driver mutations; however, its efficacy and genomic impact in EOCRC, particularly in underrepresented groups, remain poorly understood. The phosphatidylinositol 3-kinase (PI3K) pathway regulates cell growth, survival, and metabolism, and its alterations have been implicated in therapeutic resistance and adverse outcomes. Yet, the prevalence, clinical relevance, and treatment-specific associations of PI3K pathway alterations in EOCRC remain underexplored.

**Methods:** We analyzed somatic mutation and clinical data from 2,515 CRC patients (266 H/L and 2,249 Non-Hispanic White [NHW]) across publicly available genomic datasets. Patients were stratified by age at diagnosis (EOCRC <50 vs. LOCRC ≥50), ancestry (H/L vs. NHW), and FOLFOX treatment status. PI3K pathway alterations—including mutations in PIK3CA, PTEN, AKT isoforms, and regulatory genes—were identified using curated pathway definitions. Mutation prevalence was compared across groups using Fisher’s exact or chi-squared tests. AI-HOPE-PI3K, a conversational AI platform, was deployed to automate cohort construction, stratify subgroups, and perform post-hoc survival analysis.

**Results:** PI3K pathway alterations were observed across all demographic groups. In EO NHW patients treated with FOLFOX, Kaplan–Meier analysis revealed significantly reduced overall survival among those with PI3K pathway alterations (n = 124) compared with unaltered counterparts (n = 251; p = 0.0008), identifying alterations as a candidate prognostic biomarker in this subgroup. AI-guided subgroup interrogation further highlighted mutation-specific signals: INPP4B and RPTOR emerged as exploratory candidates in EO H/L patients but did not show significant treatment- or ancestry-specific enrichment upon confirmatory testing. Similarly, ancestry-stratified analysis of PIK3R2 mutations revealed comparable rates in EO H/L (1.37%) and EO NHW (1.6%) FOLFOX-treated patients (p = 1.0). Across ancestry and age groups, mutational landscape analysis revealed diverse molecular events—including missense, nonsense, splice-site, frameshift, and in-frame deletions—underscoring the heterogeneity of PI3K pathway dysregulation.

**Conclusions:** This study identifies PI3K pathway alterations as a potential prognostic marker of poor survival in EO NHW patients receiving FOLFOX and uncovers ancestry- and treatment-specific mutational differences in high-risk CRC populations. By integrating clinical, molecular, and treatment variables, the AI-HOPE and AI-HOPE-PI3K platforms enabled rapid, reproducible, and fine-grained analysis of complex datasets. These findings underscore the need for ancestry-informed molecular profiling to optimize therapeutic strategies and highlight AI-guided interrogation as a powerful tool for advancing precision oncology in underrepresented and disproportionately affected CRC populations.

## 1. Introduction

Colorectal cancer (CRC) remains the third leading cause of both cancer incidence and cancer-related mortality in the United States, despite being among the most preventable cancers through lifestyle modification, diet, and early detection strategies (1,2). While overall CRC rates in high-income countries have declined or stabilized due to screening programs, the incidence of early-onset CRC (EOCRC), defined as diagnosis before age 50, is rising at an alarming rate (1–5). By 2030, EOCRC is projected to become the leading cause of cancer-related death among individuals aged 20–49 (3–9). This increase is evident across all racial and ethnic groups but is most pronounced in Hispanic/Latino (H/L) individuals, who now represent the fastest-growing segment of the U.S. labor force and contribute ∼$4.2 trillion annually to the national economy—an amount surpassing the GDP of India, the UK, or France (1,2,6,10–12). These demographic, economic, and public health realities underscore the urgency of understanding the biological and molecular underpinnings of EOCRC in high-risk, underserved populations.

Molecular profiling studies have begun to reveal differences between EOCRC and late-onset CRC (LOCRC), including distinct mutational landscapes, LINE-1 hypomethylation, and variations in key oncogenic drivers (13–17). However, results have been inconsistent regarding tumor mutation burden, microsatellite instability (MSI), and immune checkpoint expression (15–17). Importantly, most studies are derived from predominantly homogeneous cohorts, limiting generalizability and perpetuating disparities in underrepresented populations such as H/L individuals (18–20). Our group and others have recently reported ethnicity-specific genomic alterations in CRC (21–23), but significant knowledge gaps remain, particularly regarding molecular features that may influence treatment outcomes.

The phosphatidylinositol 3-kinase (PI3K) signaling pathway is a central regulator of cell growth, survival, and metabolism, and its dysregulation is implicated in CRC tumorigenesis, metastasis, and resistance to therapy. Alterations in PI3K pathway genes—including *PIK3CA*, *PTEN*, *AKT1/2/3*, *TSC1/2*, *MTOR*, and *RICTOR*—are frequent in CRC and have been associated with chemotherapy resistance and poor prognosis in other cancers. However, the prevalence and clinical implications of these alterations in EOCRC, especially among H/L patients, remain poorly defined.

For patients with metastatic, microsatellite stable (MSS) CRC who lack actionable driver mutations, the American Society of Clinical Oncology (ASCO) recommends first-line treatment with FOLFOX—a regimen combining folinic acid, 5-fluorouracil (5-FU), and oxaliplatin (25,27). Although standard in this setting, recent evidence suggests EOCRC patients experience shorter overall survival and increased treatment-related toxicity compared to LOCRC patients receiving the same regimen (5). While some studies report better responses to 5-FU–based therapy in tumors with wild-type *TP53*, others find no consistent association (28–30). Furthermore, these studies have largely excluded or underrepresented H/L patients, leaving the relationship between ancestry, PI3K pathway alterations, and FOLFOX outcomes largely unexplored.

Recent developments in artificial intelligence (AI) have transformed how multi-omic data can be leveraged to uncover clinically actionable insights in cancer. Conversational AI agents such as AI-HOPE (31), which enable real-time, natural-language queries that dynamically integrate clinical, genomic, and treatment data. Building on this platform, the AI-HOPE-PI3K (32) module is specifically designed to interrogate PI3K pathway alterations in colorectal cancer, allowing researchers to systematically evaluate mutational patterns, co-occurring events, and treatment responses across large-scale, multi-ethnic datasets. Unlike traditional bioinformatics pipelines, which often require static coding frameworks and prolonged analytical cycles, AI-HOPE facilitates iterative and adaptive exploration of clinically relevant hypotheses, bridging computational oncology with translational medicine. In this study, the incorporation of AI-HOPE-PI3K provides a novel avenue to characterize the molecular determinants of PI3K pathway alterations in EOCRC, assess their prognostic and therapeutic implications in the context of FOLFOX treatment, and address disparities affecting disproportionately burdened populations such as H/L patients.

In this context, the present study investigates PI3K pathway alterations in EOCRC and LOCRC across H/L and Non-Hispanic White (NHW) patients, stratified by FOLFOX treatment status. By integrating genomic and clinical data from a large, multi-ethnic cohort, our goal is to identify ancestry- and age-specific PI3K pathway alterations associated with treatment exposure, thereby informing precision oncology approaches tailored to high-risk and underserved CRC populations.

## 2. Materials and Methods

### 2.1 Data Sources and Patient Selection

This analysis was conducted using somatic mutation and clinical annotation data from three publicly available colorectal cancer (CRC) cohorts accessible through the cBioPortal for Cancer Genomics: Colorectal Adenocarcinoma (TCGA, PanCancer Atlas), MSK-CHORD (Nature 2024), and GENIE BPC CRC. Only patients with confirmed primary colorectal, colon, or rectal adenocarcinoma were included. To ensure data integrity, a single primary tumor sample per patient was retained for analysis.

Patients were assigned to the Hispanic/Latino (H/L) group based on self-reported ethnicity, clinical annotations (“Hispanic or Latino,” “Spanish, NOS,” “Hispanic, NOS,” or “Latino, NOS”), or validated surname-based classification. All other patients with self-reported non-Hispanic White ancestry were categorized as NHW. Age at diagnosis was used to define EOCRC as <50 years and LOCRC as ≥50 years. The final study population consisted of 2,515 patients: 266 H/L (125 EOCRC, 141 LOCRC) and 2,249 NHW (677 EOCRC, 1,572 LOCRC).

The AI-HOPE-PI3K platform, a conversational artificial intelligence system designed for precision oncology, was employed to automate patient stratification by age, ancestry, treatment status, and PI3K pathway alteration profile. This platform streamlined cohort selection, minimized manual curation bias, and ensured reproducibility in downstream statistical and survival analyses.

### 2.2 Treatment Classification

Chemotherapy exposure data were extracted from structured drug administration records within each dataset. Patients were classified as FOLFOX-treated if they received a regimen combining folinic acid (leucovorin), fluorouracil (5-FU), and oxaliplatin within overlapping treatment timelines. Individuals without documented administration of all three agents were considered untreated with FOLFOX for the purposes of this study.

### 2.3 PI3K Pathway Alteration Definition

Genes assigned to the PI3K signaling pathway were identified using curated CRC-specific literature and pathway reference databases. The gene list included PIK3CA, PTEN, AKT1, AKT2, AKT3, MTOR, RICTOR, TSC1, TSC2, INPP4B, PIK3R1, PIK3R2, and PPP2R1A. Somatic alterations were restricted to nonsynonymous variants with potential functional impact: missense mutations, nonsense mutations, splice-site variants, frameshift insertions/deletions, in-frame deletions, and multi-hit events. Alteration burden was defined as the presence of at least one qualifying mutation in any PI3K pathway gene. The AI-HOPE-PI3K framework was further used to automatically query and classify gene-level events, enabling parallel exploration of ancestry-specific and treatment-specific mutational patterns.

### 2.4 Statistical Analysis

Comparisons of mutation prevalence between groups defined by age (EOCRC vs. LOCRC), ancestry (H/L vs. NHW), and treatment status (FOLFOX-treated vs. untreated) were performed using Fisher’s exact test or chi-squared test, depending on expected cell counts. Mutation frequencies were calculated at the gene level and across the PI3K pathway as a whole.

Mutational landscape visualizations were generated to illustrate differences in alteration type distribution (splice-site, missense, nonsense, frameshift deletion, in-frame deletion, multi-hit) across subgroups. All statistical analyses were conducted in R (version 4.3.1), with two-sided p-values <0.05 considered statistically significant.

The AI-HOPE [31] and AI-HOPE-PI3K [32] platforms were also applied to generate Kaplan–Meier survival analyses, integrating clinical annotations with pathway-specific mutation data. This AI-enabled workflow reduced manual preprocessing steps, ensured reproducibility, and allowed for rapid hypothesis testing across multi-ethnic CRC cohorts.

## 3. Results

### 3.1 Clinical and Demographic Profile of the Study Cohorts

The study included 2,515 CRC patients, comprising 266 H/L and 2,249 NHW individuals (Table 1). EOCRC (<50 years) was more common in the H/L cohort compared to NHW patients (46.9% vs. 30.1%), whereas LOCRC (≥50 years) predominated among NHW individuals (69.9% vs. 53.0%).

**Table 1.** Demographic and clinical characteristics of Hispanic/Latino (H/L) and Non-Hispanic White (NHW) colorectal cancer cohorts.

| Clinical Feature | H/L Cohort<br>n (%) | NHW Cohort<br>n (%) |
| --- | --- | --- |
| **Age Onset & Treatment** |  |  |
| Early-Onset Colorectal Cancer | 125 (46.9%) | 677 (30.1%) |
| Late-Onset Colorectal Cancer | 141 (53%) | 1572 (69.9%) |
| **Sample Type** |  |  |
| Primary Tumor | 266(100%) | 2249 (100%) |
| **Sex** |  |  |
| Male | 158 (59.4%) | 1267 (56.3%) |
| Female | 108 (40.6%) | 982 (43.7%) |
| **Sample Type** |  |  |
| Primary Tumor | 266<br>(100.0%) | 2249 (100.0%) |
| **Stage at Diagnosis** |  |  |
| Stage 1-3 | 156 (58.6%) | 1236 (55.0%) |
| Stage 4 | 108 (40.6%) | 1005 (44.7%) |
| NA | 2 (0.8%) | 8 (0.4%) |
| **Ethnicity** |  |  |
| Hispanic/Latinos (H/L) | 266 (100%) | 0 (0.0%) |
| Non-Hispanic Whites (NHW) | 0 (0.0%) | 2249 (100.0%) |

When stratified by both age and FOLFOX exposure, EOCRC patients receiving FOLFOX represented 27.4% of the H/L group versus 16.7% of the NHW group. LOCRC patients treated with FOLFOX accounted for 34.2% of H/L and 40.9% of NHW patients. Among those not treated with FOLFOX, EOCRC cases comprised 19.5% of H/L and 13.4% of NHW patients, while LOCRC made up 18.8% and 29.0%, respectively.

All analyzed samples were primary tumor specimens. Sex distribution was comparable across groups, with males representing 59.4% of H/L and 56.3% of NHW patients. Stage at diagnosis showed similar trends between cohorts: stage I–III disease was present in 58.6% of H/L and 55.0% of NHW patients, while stage IV disease occurred in 40.6% and 44.7%, respectively.

By design, all patients in the H/L cohort self-identified as Hispanic or Latino, and all patients in the NHW cohort self-identified as NHW, ensuring clear ancestry stratification for subsequent genomic analyses.

These baseline characteristics demonstrate both shared and distinct features between cohorts, notably the higher proportion of EOCRC and FOLFOX-treated young patients in the H/L group, providing a rationale for ancestry- and treatment-specific PI3K pathway analyses.

### 3.2 Comparative Genomic Analysis by Age and Ancestry

Comparative analysis of PI3K pathway gene alterations between EOCRC and LOCRC in H/L patients revealed distinct treatment-associated patterns (Tables 2a, S1 and S2). In EOCRC, INPP4B mutations were present in 9.6% of untreated patients but absent in those receiving FOLFOX (*p* = 0.011). PPP2R1A mutations followed a similar trend, occurring in 7.7% of untreated patients and none of the treated cases (*p* = 0.028). In LOCRC, RICTOR mutations were observed in 8.0% of untreated patients but absent among those treated with FOLFOX (*p* = 0.015).

Among NHW patients (Table 2b, S3 and S4), several PI3K pathway genes demonstrated age- and treatment-related differences. In EOCRC, AKT3 mutations were significantly less frequent in treated patients (0.8%) compared to untreated patients (3.0%, *p* = 0.041), while AKT2 mutations occurred at low frequencies in both treated (0.5%) and untreated (0.7%) groups (*p* = 1.00). In LOCRC, MTOR mutations were significantly reduced in the treated group (4.5%) versus untreated (7.8%, *p* = 0.007), and AKT2 mutations approached significance (0.7% vs. 1.8%, *p* = 0.053). No significant differences in AKT3 mutation rates were detected in LOCRC (1.0% vs. 1.8%, *p* = 0.216).

**Table 2.**
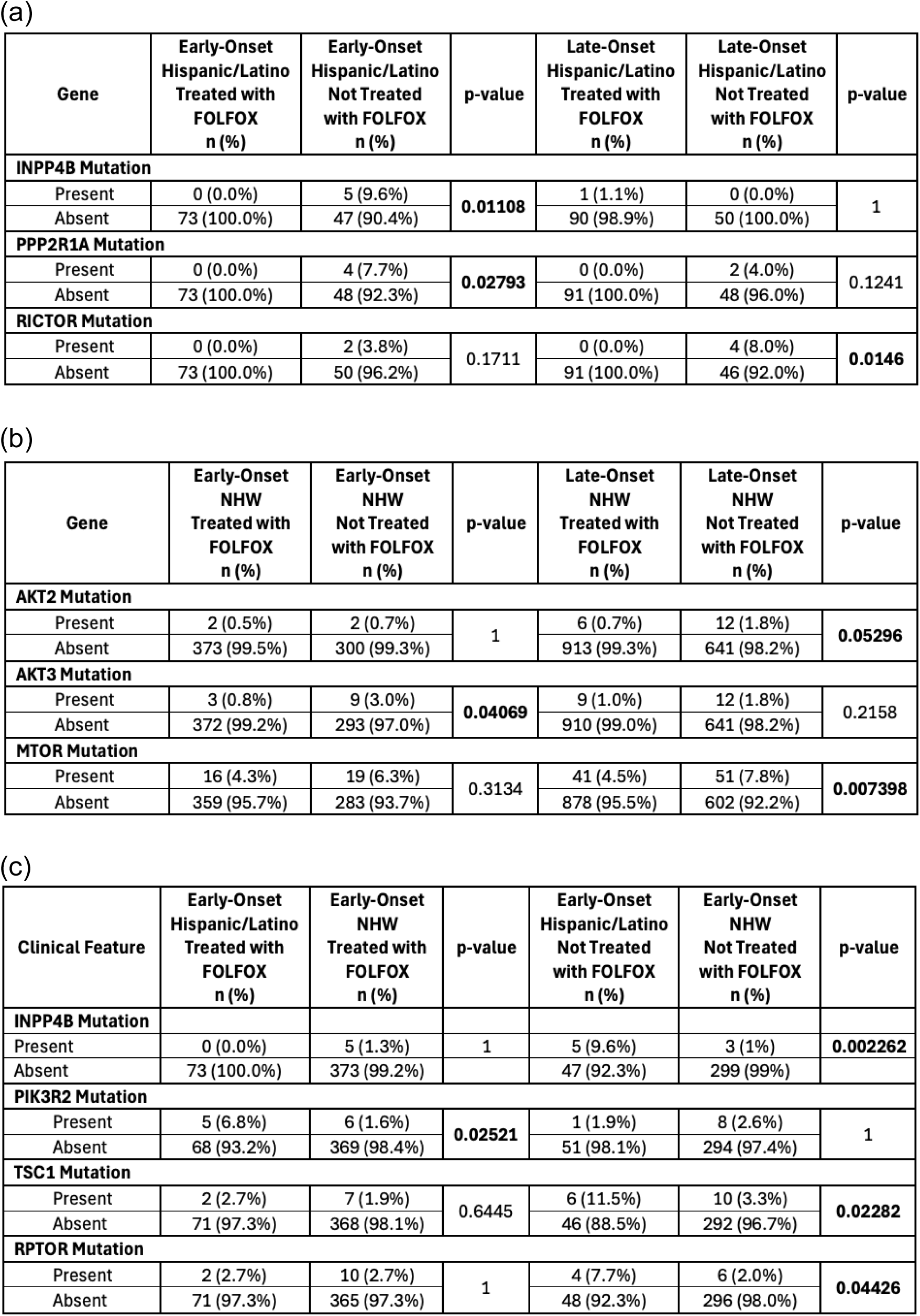
Comparative analysis of PI3K pathway gene alterations in colorectal cancer (CRC) subgroups stratified by age, ancestry, and FOLFOX treatment status. This table presents subgroup-specific mutation frequencies for select PI3K pathway genes across: (a) Early-Onset (EOCRC) vs. Late-Onset (LOCRC) Hispanic/Latino (H/L) patients; (b) EOCRC vs. LOCRC Non-Hispanic White (NHW) patients; and (c) EOCRC comparisons between H/L and NHW cohorts. Results are shown separately for patients treated with FOLFOX and those without FOLFOX exposure, highlighting statistically significant differences in mutation prevalence.

| Gene | Early-Onset<br>Hispanic/Latino<br>Treated with<br>FOLFOX<br>n (%) | Early-Onset<br>Hispanic/Latino<br>Not Treated<br>with FOLFOX<br>n (%) | p-value | Late-Onset<br>Hispanic/Latino<br>Treated with<br>FOLFOX<br>n (%) | Late-Onset<br>Hispanic/Latino<br>Not Treated<br>with FOLFOX<br>n (%) | p-value |
| --- | --- | --- | --- | --- | --- | --- |
| INPP4B Mutation |  |  |  |  |  |  |
| Present | 0 (0.0%) | 5 (9.6%) | 0.01108 | 1 (1.1%) | 0 (0.0%) | 1 |
| Absent | 73 (100.0%) | 47 (90.4%) |  | 90 (98.9%) | 50 (100.0%) |  |
| PPP2R1A Mutation |  |  |  |  |  |  |
| Present | 0 (0.0%) | 4 (7.7%) | 0.02793 | 0 (0.0%) | 2 (4.0%) | 0.1241 |
| Absent | 73 (100.0%) | 48 (92.3%) |  | 91 (100.0%) | 48 (96.0%) |  |
| RICTOR Mutation |  |  |  |  |  |  |
| Present | 0 (0.0%) | 2 (3.8%) | 0.1711 | 0 (0.0%) | 4 (8.0%) | 0.0146 |
| Absent | 73 (100.0%) | 50 (96.2%) |  | 91 (100.0%) | 46 (92.0%) |  |

(b)
| Gene | Early-Onset<br>NHW<br>Treated with<br>FOLFOX<br>n (%) | Early-Onset<br>NHW<br>Not Treated<br>with FOLFOX<br>n (%) | p-value | Late-Onset<br>NHW<br>Treated with<br>FOLFOX<br>n (%) | Late-Onset<br>NHW<br>Not Treated<br>with FOLFOX<br>n (%) | p-value |
| --- | --- | --- | --- | --- | --- | --- |
| AKT2 Mutation |  |  |  |  |  |  |
| Present | 2 (0.5%) | 2 (0.7%) | 1 | 6 (0.7%) | 12 (1.8%) | 0.05296 |
| Absent | 373 (99.5%) | 300 (99.3%) |  | 913 (99.3%) | 641 (98.2%) |  |
| AKT3 Mutation |  |  |  |  |  |  |
| Present | 3 (0.8%) | 9 (3.0%) | 0.04069 | 9 (1.0%) | 12 (1.8%) | 0.2158 |
| Absent | 372 (99.2%) | 293 (97.0%) |  | 910 (99.0%) | 641 (98.2%) |  |
| MTOR Mutation |  |  |  |  |  |  |
| Present | 16 (4.3%) | 19 (6.3%) | 0.3134 | 41 (4.5%) | 51 (7.8%) | 0.007398 |
| Absent | 359 (95.7%) | 283 (93.7%) |  | 878 (95.5%) | 602 (92.2%) |  |

(c)
| Clinical Feature | Early-Onset<br>Hispanic/Latino<br>Treated with<br>FOLFOX<br>n (%) | Early-Onset<br>NHW<br>Treated with<br>FOLFOX<br>n (%) | p-value | Early-Onset<br>Hispanic/Latino<br>Not Treated<br>with FOLFOX<br>n (%) | Early-Onset<br>NHW<br>Not Treated<br>with FOLFOX<br>n (%) | p-value |
| --- | --- | --- | --- | --- | --- | --- |
| INPP4B Mutation |  |  |  |  |  |  |
| Present | 0 (0.0%) | 5 (1.3%) | 1 | 5 (9.6%) | 3 (1%) | 0.002262 |
| Absent | 73 (100.0%) | 373 (99.2%) |  | 47 (92.3%) | 299 (99%) |  |
| PIK3R2 Mutation |  |  |  |  |  |  |
| Present | 5 (6.8%) | 6 (1.6%) | 0.02521 | 1 (1.9%) | 8 (2.6%) | 1 |
| Absent | 68 (93.2%) | 369 (98.4%) |  | 51 (98.1%) | 294 (97.4%) |  |
| TSC1 Mutation |  |  |  |  |  |  |
| Present | 2 (2.7%) | 7 (1.9%) | 0.6445 | 6 (11.5%) | 10 (3.3%) | 0.02282 |
| Absent | 71 (97.3%) | 368 (98.1%) |  | 46 (88.5%) | 292 (96.7%) |  |
| RPTOR Mutation |  |  |  |  |  |  |
| Present | 2 (2.7%) | 10 (2.7%) | 1 | 4 (7.7%) | 6 (2.0%) | 0.04426 |
| Absent | 71 (97.3%) | 365 (97.3%) |  | 48 (92.3%) | 296 (98.0%) |  |

Direct ancestry comparisons in EOCRC (Table 2c, S5 and S6) revealed several ancestry- and treatment-specific differences in PI3K pathway alterations. Among FOLFOX-treated patients, PIK3R2 mutations were significantly more frequent in H/L patients than in NHW patients (6.8% vs. 1.6%, *p* = 0.025), while mutation rates for INPP4B, TSC1, and RPTOR were comparable between groups. In the untreated EOCRC subgroup, multiple genes showed significant enrichment in H/L patients. INPP4B mutations were observed in 9.6% of H/L patients compared to 1.0% of NHW patients (*p* = 0.002). TSC1 mutations were also more common in H/L patients (11.5% vs. 3.3%, *p* = 0.023), as were RPTOR mutations (7.7% vs. 2.0%, *p* = 0.044). No significant ancestry-based difference was detected for PIK3R2 in untreated EOCRC (1.9% vs. 2.6%, *p* = 1.00).

### 3.3 Prevalence of PI3K Pathway Alterations by Age, Ancestry, and FOLFOX Treatment Status

An integrated analysis of PI3K pathway alterations revealed a moderate prevalence across all colorectal cancer (CRC) subgroups, with no statistically significant differences by age of onset, ancestry, or FOLFOX treatment status.

In the H/L EOCRC subgroup (Table 3a), PI3K alterations were detected in 35.6% of FOLFOX-treated patients and 44.2% of untreated patients (*p* = 0.43). Among LOCRC patients, alterations were observed in 44.0% of treated and 36.0% of untreated cases (*p* = 0.46). While the proportion without PI3K alterations was higher in treated EOCRC (64.4% vs. 55.8%), the opposite pattern occurred in LOCRC (56.0% vs. 64.0%). Overall, PI3K pathway dysregulation was common in H/L CRC, but variation by age or chemotherapy exposure was modest.

**Table 3.** Frequency of PI3K Pathway Alterations in Hispanic/Latino and Non-Hispanic White Colorectal Cancer Patients Stratified by Age of Onset and FOLFOX Treatment Status. This table summarizes the mutation frequencies of key genes involved in the PI3K signaling pathway—PTEN, PIK3R1, PIK3R2, PIK3R3, PIK3CA, INPP4B, AKT1, AKT2, AKT3, PPP2R1A, TSC1, TSC2, STK11, RHEB, RICTOR, MTOR, and RPTOR—among colorectal cancer (CRC) patients, stratified by: (3a) early-onset (EOCRC) vs. late-onset (LOCRC) and FOLFOX treatment status within the Hispanic/Latino (H/L) cohort; (3b) FOLFOX-treated vs. untreated patients within EOCRC and LOCRC subgroups of Non-Hispanic White (NHW) patients; (3c) EOCRC H/L vs. NHW patients by FOLFOX treatment status; and (3d) LOCRC H/L vs. NHW patients by FOLFOX treatment status. Statistically significant differences (p < 0.05, Chi-square or Fisher’s exact test) are indicated with asterisks. This stratified analysis highlights potential interactions between age, ancestry, chemotherapy exposure, and PI3K pathway dysregulation.

| Pathway Alterations | Early-Onset<br>Hispanic/Latino<br>Treated with<br>FOLFOX<br>n (%) | Early-Onset<br>Hispanic/Latino<br>Not Treated<br>with FOLFOX<br>n (%) | p-value | Late-Onset<br>Hispanic/Latino<br>Treated with<br>FOLFOX<br>n (%) | Late-Onset<br>Hispanic/Latino<br>Not Treated<br>with FOLFOX<br>n (%) | p-value |
| --- | --- | --- | --- | --- | --- | --- |
| PI3K Alterations Present | 26 (35.6%) | 23 (44.2%) | 0.4316 | 40 (44.0%) | 18 (36.0%) | 0.4596 |
| PI3K Alterations Absent | 47 (64.4%) | 29 (55.8%) |  | 51 (56.0%) | 32 (64.0%) |  |

(b)
| Pathway Alterations | Early-Onset<br>NHW<br>Treated with<br>FOLFOX<br>n (%) | Early-Onset<br>NHW<br>Not Treated<br>with FOLFOX<br>n (%) | p-value | Late-Onset<br>NHW<br>Treated with<br>FOLFOX<br>n (%) | Late-Onset<br>NHW<br>Not Treated<br>with FOLFOX<br>n (%) | p-value |
| --- | --- | --- | --- | --- | --- | --- |
| PI3K Alterations Present | 124 (33.1%) | 105 (34.8%) | 0.7014 | 336 (36.6%) | 266 (40.7%) | 0.1042 |
| PI3K Alterations Absent | 251 (66.9%) | 197 (65.2%) |  | 583 (63.4%) | 387 (59.3%) |  |

(c)
| Pathway Alterations | Early-Onset<br>Hispanic/Latino<br>Treated with<br>FOLFOX<br>n (%) | Early-Onset<br>NHW<br>Treated with<br>FOLFOX<br>n (%) | p-value | Early-Onset<br>Hispanic/Latino<br>Not Treated<br>with FOLFOX<br>n (%) | Early-Onset<br>NHW<br>Not Treated<br>with<br>FOLFOX<br>n (%) | p-value |
| --- | --- | --- | --- | --- | --- | --- |
| PI3K Alterations Present | 26 (35.6%) | 124 (33.1%) | 0.7743 | 23 (44.2%) | 105 (34.8%) | 0.2479 |
| PI3K Alterations Absent | 47 (64.4%) | 251 (66.9%) |  | 29 (55.8%) | 197 (65.2%) |  |

(d)
| Pathway Alterations | Late-Onset<br>Hispanic/Latino<br>Treated with<br>FOLFOX<br>n (%) | Late-Onset<br>NHW<br>Treated with<br>FOLFOX<br>n (%) | p-value | Late-Onset<br>Hispanic/Latino<br>Not Treated<br>with FOLFOX<br>n (%) | Late-Onset<br>NHW<br>Not Treated<br>with<br>FOLFOX<br>n (%) | p-value |
| --- | --- | --- | --- | --- | --- | --- |
| PI3K Alterations Present | 40 (44.0%) | 336 (36.6%) | 0.2012 | 18 (36.0%) | 266 (40.7%) | 0.6114 |
| PI3K Alterations Absent | 51 (56.0%) | 583 (63.4%) |  | 32 (64.0%) | 387 (59.3%) |  |

In the NHW cohort (Table 3b), prevalence was similar: 33.1% of FOLFOX-treated and 34.8% of untreated EOCRC patients harbored PI3K alterations (*p* = 0.7014). In LOCRC, alterations were present in 36.6% of treated and 40.7% of untreated patients (*p* = 0.1042). Across all NHW subgroups, PI3K alterations occurred in roughly one-third to two-fifths of patients, with the majority (59.3–66.9%) lacking such alterations, indicating limited impact of FOLFOX on overall pathway mutational burden.

Direct ancestry comparisons within EOCRC (Table 3c) showed no significant differences. Among FOLFOX-treated patients, alterations were present in 35.6% of H/L and 33.1% of NHW patients (*p* = 0.7743). In the untreated subgroup, the prevalence was 44.2% in H/L and 34.8% in NHW patients (*p* = 0.2479). The proportion without PI3K alterations remained high in both groups (55.8–66.9%).

Similarly, in LOCRC (Table 3d), PI3K alterations occurred in 44.0% of treated H/L patients and 36.6% of treated NHW patients (*p* = 0.2012). Among untreated patients, prevalence was 36.0% in H/L and 40.7% in NHW (*p* = 0.6114).

Collectively, these results demonstrate that PI3K pathway alterations are moderately prevalent across CRC subgroups and occur at comparable frequencies regardless of age of onset, ancestry, or FOLFOX exposure. While specific gene-level differences may exist, the overall burden of PI3K dysregulation appears to be a shared molecular feature in both H/L and NHW populations.

### 3.4 Mutational Landscape

#### PI3K Pathway Alterations in Early-Onset Hispanic/Latino CRC

To assess the distribution of PI3K pathway alterations in EOCRC among H/L patients, we analyzed somatic mutations in 17 key pathway genes using oncoplot visualization (Figure 1A, Table S7). Of 123 EO H/L CRC cases, 49 (39.8%) harbored at least one alteration in a PI3K pathway gene.

**Figure 1.**
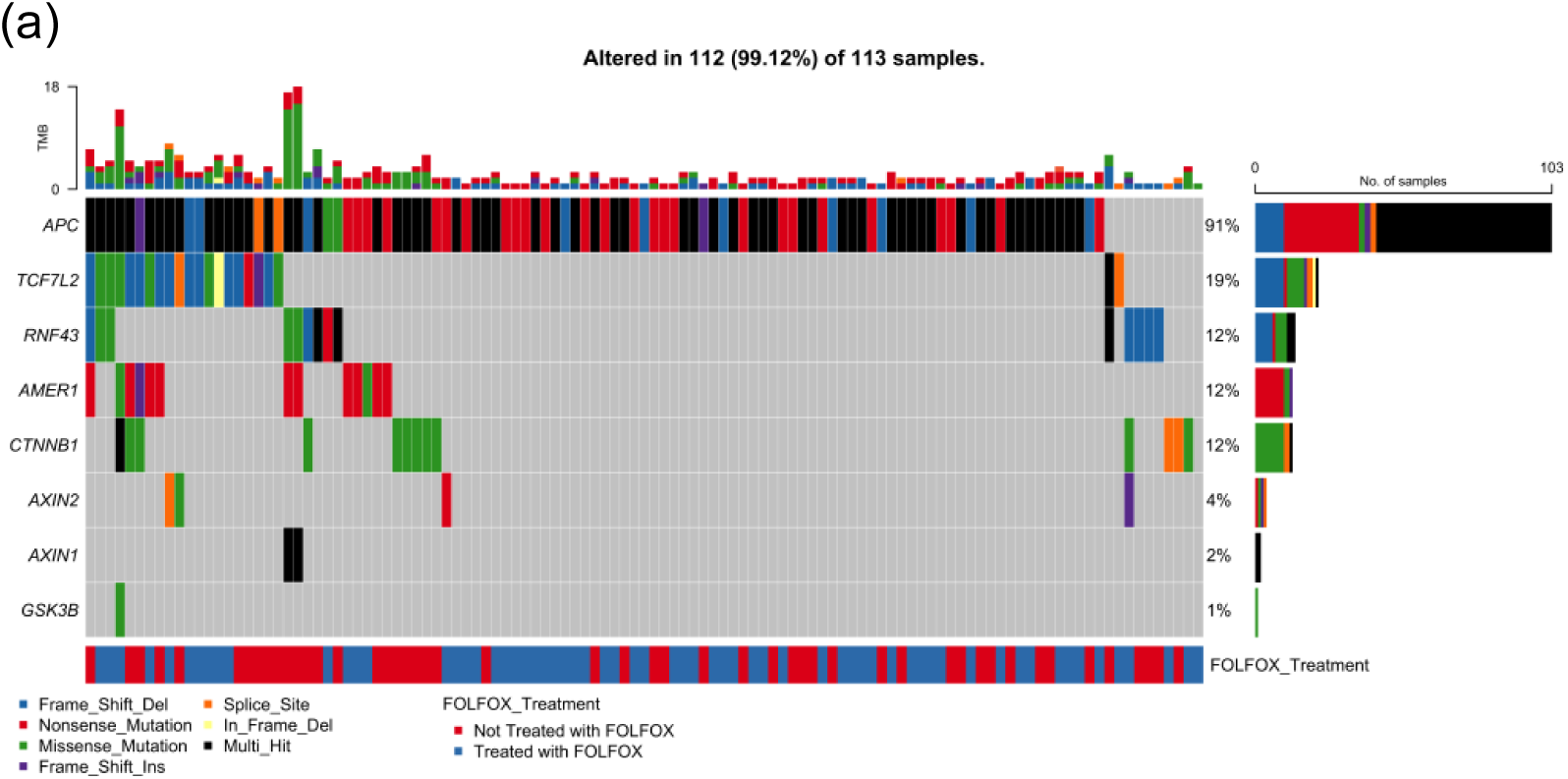
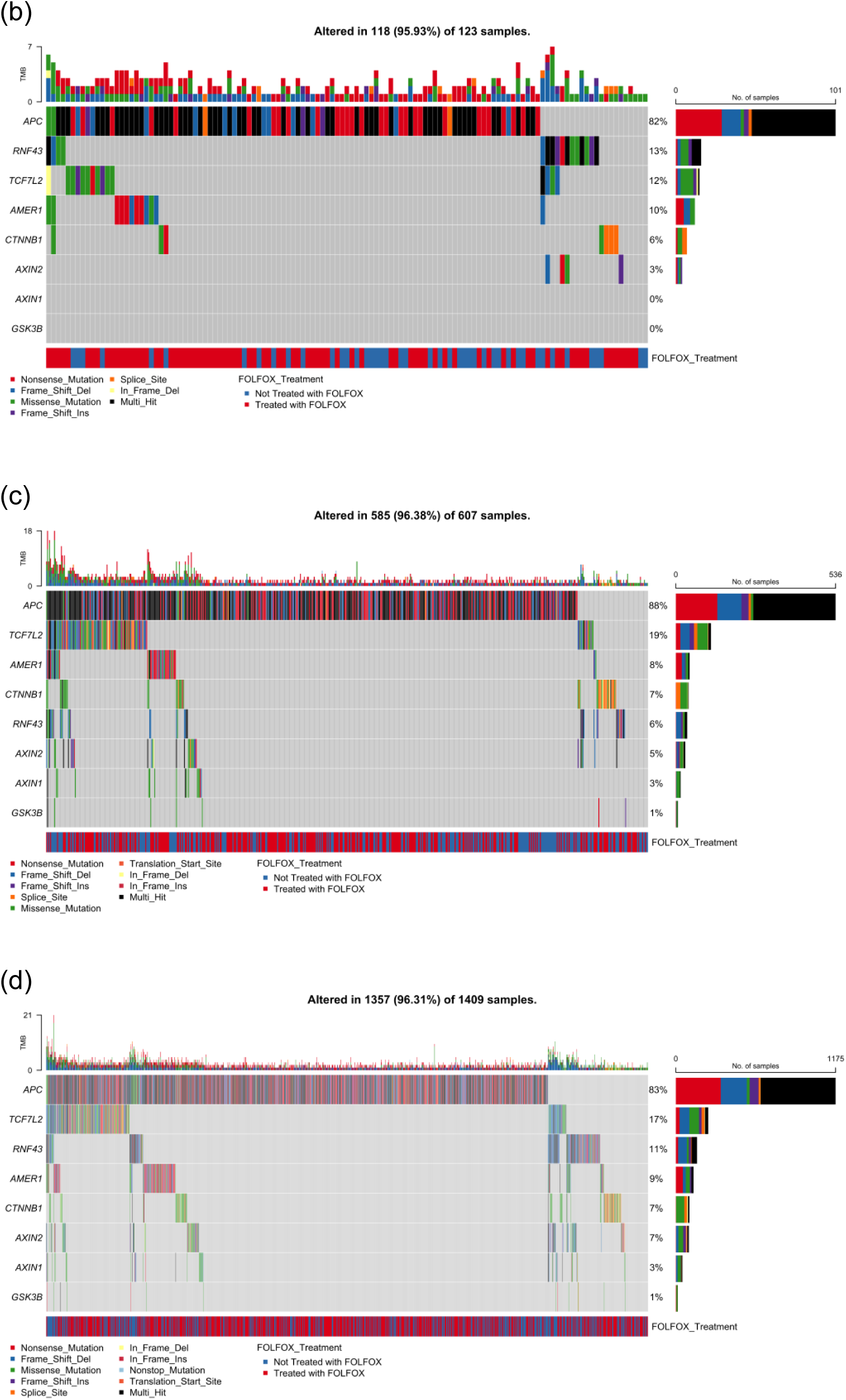
Somatic mutation landscape of PI3K pathway genes in colorectal cancer (CRC) stratified by age, ancestry, and FOLFOX treatment status. Oncoplots displaying gene-level mutation profiles of PI3K pathway components in colorectal cancer, stratified by age of onset (early vs. late) and ancestry (Hispanic/Latino vs. Non-Hispanic White). Panels show mutation types, tumor mutational burden (TMB), and FOLFOX treatment status across: (a) 123 early-onset Hispanic/Latino (H/L) patients, (b) 140 late-onset H/L patients, (c) 660 early-onset Non-Hispanic White (NHW) patients, and (d) 1,537 late-onset NHW patients. Across all subgroups, PIK3CA is the most frequently mutated gene, with a predominance of missense variants, followed by alterations in PTEN, MTOR, and other pathway regulators such as TSC1/2, PIK3R1, and RICTOR. The data reveal pervasive disruption of PI3K-AKT-mTOR signaling in CRC, with variation in alteration frequency and spectrum by age and ancestry, and no distinct clustering associated with FOLFOX treatment.

PIK3CA was the most frequently mutated gene (22%), predominantly with missense variants, followed by PTEN (10%), which exhibited a diverse mutation spectrum including missense, nonsense, and truncating frame shift events. Other recurrently altered genes included TSC1 (7%), TSC2 (7%), and MTOR (7%). Additional affected genes—PIK3R2 (5%), AKT1 (5%), RPTOR (5%), and INPP4B (5%)—were primarily altered through missense or truncating mutations. Less common events were detected in AKT3 (4%), PIK3R1 (3%), PPP2R1A (3%), and PIK3R3 (2%). No alterations were observed in AKT2, STK11, or RHEB.

Tumor mutational burden (TMB) was generally low, though a subset displayed hypermutated profiles suggestive of mismatch repair deficiency. FOLFOX treatment status did not show distinct clustering of mutation patterns. These results identify PIK3CA and PTEN as primary drivers, with additional alterations distributed across multiple regulatory and effector nodes of the pathway, highlighting therapeutic targets for this high-risk population.

#### PI3K Pathway Alterations in Late-Onset Hispanic/Latino CRC

In 140 LOCRC H/L cases, 58 (41.4%) carried at least one PI3K pathway alteration (Figure 1B, Table S7). PIK3CA was again the most frequently mutated gene (24%), followed by PTEN, TSC2, and MTOR (5% each). PIK3R1, PIK3R2, AKT1, AKT3, and TSC1 each occurred in 3% of cases, while PIK3R3, AKT2, PPP2R1A, and INPP4B were altered in ∼1% of cases. No mutations were detected in STK11, RHEB, RICTOR, or RPTOR.

PIK3CA alterations were primarily missense, whereas PTEN carried both truncating and missense variants. MTOR and TSC2 were enriched for missense mutations, with occasional in-frame insertions or deletions. PI3K regulatory subunits (PIK3R1–3) exhibited both missense and truncating events.

TMB was generally low, with occasional hypermutated outliers. No clear enrichment or clustering of mutations was observed by FOLFOX treatment status. These findings reinforce PIK3CA as the dominant driver in LO H/L CRC, with additional contributions from tumor suppressors and mTOR-associated genes.

#### PI3K Pathway Alterations in Early-Onset Non-Hispanic White CRC

Among 660 EOCRC cases in NHW patients, 229 (34.7%) exhibited at least one PI3K pathway mutation (Figure 1C, Table S7). PIK3CA (21%) was the most recurrently altered, with a predominance of missense and multi-hit variants. Other frequent alterations included PIK3R1 (7%), PTEN (6%), MTOR (5%), and TSC2 (4%). PPP2R1A and RICTOR each occurred in 3% of cases, while TSC1, PIK3R2, AKT1, and AKT3 were found in ∼2%. Less common events (<1%) involved PIK3R3, INPP4B, STK11, and AKT2; RHEB mutations were absent.

TMB distribution was low overall, with occasional hypermutated profiles. No distinct mutation patterns were linked to FOLFOX exposure. This cohort demonstrates recurrent alterations spanning both canonical PI3K-AKT signaling and mTOR regulation, supporting exploration of targeted therapeutic strategies.

#### PI3K Pathway Alterations in Late-Onset Non-Hispanic White CRC

In 1,537 LOCRC cases from NHW patients, 602 (39.2%) harbored at least one PI3K pathway alteration (Figure 1D, Table S13). PIK3CA mutations (22%) dominated, primarily as missense variants, followed by MTOR (6%) and PTEN (6%). PIK3R1 was altered in 5% of cases, often with truncating or splice site variants. TSC2 (4%), RICTOR (3%), PPP2R1A (3%), RPTOR (3%), and AKT1 (2%) were also recurrently altered. AKT3, PIK3R2, INPP4B, STK11, and AKT2 mutations occurred in ≤2% of samples; PIK3R3 and RHEB were unaltered.

TMB remained low in most cases, with sporadic hypermutated tumors. FOLFOX treatment status showed no consistent association with specific mutation patterns. The breadth of altered nodes, from upstream regulators to downstream effectors, underscores the multiple routes through which PI3K signaling can be dysregulated in LOCRC.

### 3.5 AI-Enabled Data Interrogation and Pre-Statistical Insights

The AI-HOPE-PI3K platform was deployed for targeted, post-hoc interrogation of integrated colorectal cancer datasets, using natural language–driven queries to rapidly identify survival-associated genomic features. This AI-assisted workflow facilitated hypothesis generation and streamlined subsequent statistical validation, with an initial focus on EOCRC NHW patients treated with FOLFOX.

In this cohort, AI-guided queries highlighted a potential association between PI3K pathway alterations and reduced overall survival. Kaplan–Meier analysis (Figure S1) comparing 124 patients with PI3K pathway alterations to 251 without such alterations confirmed a significant survival disadvantage for the altered group (log-rank p = 0.0008). The altered group exhibited an early and persistent decline in survival probability within the first ∼40 months, with divergence maintained over the follow-up period. Narrower confidence intervals and a consistent gap between curves reinforced the robustness of the association, suggesting PI3K pathway alterations as a potential prognostic biomarker for poorer outcomes in EO NHW patients receiving FOLFOX.

AI-driven subgroup interrogation next examined INPP4B mutation prevalence in early-onset H/L CRC patients by treatment status. A case cohort of FOLFOX-treated EO H/L patients (n = 73) was compared with an untreated EO H/L control group (n = 52). Fisher’s exact test revealed no significant difference (p = 0.864; OR = 0.0, 95% CI: 0.012–10.161), with mutations present in 0.68% and 1.92% of patients, respectively (Figure S2). These results indicate that while AI-based scanning flagged INPP4B for potential variability, confirmatory testing did not support a treatment-specific enrichment pattern.

The platform also evaluated RPTOR mutation prevalence between non-FOLFOX–treated EO H/L CRC patients (n = 73) and matched EO NHW controls (n = 375). Fisher’s exact test showed no significant difference (p = 0.328; OR = 0.0, 95% CI: 0.014–4.328), with mutations present in 0.68% of cases and 2.67% of controls (Figure S3). While RPTOR emerged during exploratory PI3K pathway interrogation, these results did not support a significant ancestry-specific enrichment in this subset.

In ancestry-stratified analyses of PIK3R2 mutations among FOLFOX-treated EO H/L and EO NHW patients, prevalence was 1.37% (n = 73) and 1.6% (n = 375), respectively. Fisher’s exact test indicated no significant difference (p = 1.0; OR = 0.854, 95% CI: 0.101– 7.202) (Figures S4–S5). Although mutation rates were low, these results demonstrate the platform’s capacity to rapidly stratify large-scale multi-omics datasets by complex demographic, clinical, and treatment variables—an essential feature for pathway-specific, high-risk subgroup analyses.

Collectively, these exploratory outputs informed targeted subgroup comparisons for formal statistical testing. The AI system executed automated filtering, cohort construction, and mutation frequency tabulation, integrating clinical, molecular, and treatment parameters. This reduced manual data handling, ensured reproducibility, and accelerated the transition from hypothesis generation to statistical validation. By enabling rapid, reproducible, and fine-grained interrogation of complex datasets, the AI-HOPE-PI3K platform supports discovery of ancestry- and treatment-specific molecular patterns within the PI3K signaling axis, advancing precision oncology research in high-risk colorectal cancer populations.

## 4. Discussion

The rising incidence of early-onset colorectal cancer (EOCRC), particularly in H/L populations, continues to underscore the need for ancestry- and treatment-informed molecular analyses. This study represents one of the largest comparative investigations of PI3K pathway alterations across EOCRC and LOCRC, integrating ancestry and chemotherapy exposure as critical modifiers of genomic architecture. By applying both traditional statistical approaches and AI-enabled interrogation, we demonstrate that the prevalence, spectrum, and treatment associations of PI3K pathway alterations differ substantially between H/L and NHW patients, highlighting the heterogeneity of CRC biology across high-risk groups.

### Ancestry- and Age-Specific Alteration Patterns

Our findings confirm that PI3K pathway alterations are common across all CRC subgroups, with PIK3CA emerging as the most recurrently mutated gene, followed by PTEN, MTOR, and regulatory subunits such as PIK3R1 and PIK3R2. Importantly, ancestry-specific differences were evident. Untreated EO H/L patients harbored significantly higher frequencies of TSC1, RPTOR, and INPP4B mutations compared with untreated EO NHW patients, suggesting distinct mechanisms of PI3K pathway dysregulation in younger H/L cases. These differences may reflect ancestry-related variation in environmental exposures, germline susceptibility, or tumor evolution trajectories, consistent with prior studies reporting ethnicity-specific genomic signatures in CRC. The enrichment of TSC1 and RPTOR mutations in untreated H/L patients, for instance, may implicate alternative mTOR complex signaling vulnerabilities unique to this population.

### Impact of FOLFOX Chemotherapy on PI3K Alterations

Chemotherapy exposure was associated with selective differences in the prevalence of PI3K pathway mutations. In EO H/L patients, FOLFOX-treated tumors exhibited striking reductions in INPP4B and PPP2R1A mutations compared to untreated counterparts, whereas LO H/L tumors showed loss of RICTOR alterations following treatment. Among NHW patients, FOLFOX-treated EOCRC cases demonstrated reduced AKT3 mutation frequencies, while LOCRC cases exhibited lower rates of MTOR and AKT2 alterations. Collectively, these findings suggest that FOLFOX may exert selective pressure on specific PI3K pathway nodes, potentially eliminating sensitive subclones or enriching resistant lineages. While these associations cannot establish causality in the absence of longitudinal sampling, they raise important questions regarding whether PI3K pathway alterations confer differential treatment resistance—or whether chemotherapy itself reshapes the PI3K mutational landscape.

### Cross-Population Insights and Clinical Implications

Perhaps the most notable ancestry-specific observation was the higher frequency of PIK3R2 mutations in FOLFOX-treated EO H/L patients compared to their NHW counterparts. Given the role of PIK3R2 in regulating PI3K complex stability, this enrichment may reflect an ancestry-linked vulnerability to chemotherapy or a context-specific resistance mechanism. The consistent presence of PIK3CA mutations across all groups reinforces its role as a foundational driver of CRC biology, while the variable involvement of ancillary regulators such as INPP4B, TSC1, and RICTOR underscores the complexity of PI3K pathway disruption. These findings point toward the need for ancestry-aware biomarker development and tailored therapeutic strategies, particularly in underserved populations disproportionately affected by EOCRC.

### AI-Guided Precision Oncology

The integration of the AI-HOPE-PI3K platform provided unique advantages for hypothesis generation and subgroup interrogation. AI-guided queries rapidly identified survival-associated PI3K alterations in EO NHW patients treated with FOLFOX, which were subsequently validated by Kaplan–Meier analysis, revealing significantly worse overall survival among altered cases. Moreover, the system enabled rapid cross-comparisons of low-frequency alterations such as INPP4B, RPTOR, and PIK3R2 across ancestry and treatment-defined subgroups, streamlining the transition from exploratory insights to statistical validation. This approach exemplifies how conversational AI can overcome traditional bioinformatics bottlenecks by dynamically integrating molecular, clinical, and treatment data to support precision oncology research.

### Limitations and Future Directions

Several limitations warrant consideration. The retrospective use of publicly available datasets introduces potential biases in treatment annotation and may underrepresent chemotherapy-specific sequencing data. The relatively small sample sizes within some ancestry- and treatment-defined subgroups limit the statistical power for detecting low-frequency events. Furthermore, survival analyses were constrained by incomplete clinical annotation for non-NHW cohorts, precluding definitive conclusions regarding the prognostic impact of PI3K alterations in H/L patients. Prospective, ancestry-diverse studies with harmonized clinical, molecular, and treatment annotation will be critical to validate and extend these findings.

Functional studies using high-definition genomic and spatial biology techniques [33] are urgently needed to disentangle whether the differential prevalence of PI3K pathway alterations in treated versus untreated cohorts reflects true chemotherapy-driven clonal selection or pre-existing biological heterogeneity. Traditional bulk sequencing captures aggregate mutational patterns but lacks the spatial resolution to resolve clonal architecture within the tumor microenvironment (TME). Emerging technologies such as spatial transcriptomics, multiplexed imaging, and spatial proteomics [34] now enable in situ mapping of PI3K-altered subclones, immune infiltration, and stromal interactions at single-cell resolution [35]. These approaches can illuminate whether selective loss of alterations such as *INPP4B* or *RICTOR* following FOLFOX represents direct therapeutic elimination of sensitive clones or instead reflects the outgrowth of resistant populations already present at low frequency. Parallel advances in single-cell DNA/RNA sequencing further support the reconstruction of lineage trajectories under chemotherapy pressure, offering mechanistic insights into evolutionary bottlenecks and adaptive resistance. Importantly, applying these methods in ancestry-diverse CRC cohorts is essential, as spatial context and clonal dynamics may vary by genetic ancestry, influencing both mutational landscapes and treatment responses. Ultimately, combining AI-guided patient stratification with spatial and single-cell functional validation will help clarify the causal role of PI3K alterations in modulating chemotherapy outcomes and may identify new biomarkers or therapeutic targets to improve survival in disproportionately affected EOCRC populations.

## 5. Conclusions

This study reveals that PI3K pathway alterations are moderately prevalent in CRC across ancestry and age groups, but with significant heterogeneity in gene-level patterns and chemotherapy associations. H/L patients, particularly those with EOCRC, exhibited distinct PI3K pathway profiles compared to NHW counterparts, underscoring the importance of ancestry-informed molecular profiling. The use of AI-enabled interrogation further highlighted PI3K alterations as potential prognostic biomarkers in EOCRC and demonstrated the power of conversational AI in precision oncology. Together, these findings emphasize that integrating ancestry, treatment context, and AI-driven discovery is essential to developing equitable and effective therapeutic strategies for high-risk CRC populations.

## Data Availability

All data used in the present study is publicly available at https://www.cbioportal.org/ and https://genie.cbioportal.org. The datasets used in our study were aggregated/summary data, and no individual-level data were used. Additional data can be provided upon reasonable request to the authors.

## Supplementary Materials

**Table S1.** EO HL Treated with FOLFOX v EO HL Not Treated with FOLFOX.

| Gene | Early-Onset Hispanic/Latino<br>Treated with FOLFOX<br>n (%) | Early-Onset Hispanic/Latino<br>Not Treated with FOLFOX<br>n (%) | p-value |
| --- | --- | --- | --- |
| PTEN Mutation |  |  |  |
| Present | 4 (5.5%) | 8 (15.4%) | 0.07366 |
| Absent | 69 (94.5%) | 44 (84.6%) |  |
| PIK3R1 Mutation |  |  |  |
| Present | 1 (1.4%) | 3 (5.8%) | 0.3067 |
| Absent | 72 (98.6%) | 49 (94.2%) |  |
| PIK3R2 Mutation |  |  |  |
| Present | 5 (6.8%) | 1 (1.9%) | 0.3993 |
| Absent | 68 (93.2%) | 51 (98.1%) |  |
| PIK3R3 Mutation |  |  |  |
| Present | 1 (1.4%) | 1 (1.9%) | 1 |
| Absent | 72 (98.6%) | 51 (98.1%) |  |
| PIK3CA Mutation |  |  |  |
| Present | 14 (19.2%) | 13 (25.0%) | 0.5761 |
| Absent | 59 (80.8%) | 39 (75.0%) |  |
| INPP4B Mutation |  |  |  |
| Present | 0 (0.0%) | 5 (9.6%) | 0.01108 |
| Absent | 73 (100.0%) | 47 (90.4%) |  |
| AKT1 Mutation |  |  |  |
| Present | 4 (5.5%) | 2 (3.8%) | 1 |
| Absent | 69 (94.5%) | 50 (96.2%) |  |
| AKT2 Mutation |  |  |  |
| Present | 0 (0.0%) | 0 (0.0%) | 1 |
| Absent | 73 (100.0%) | 52 (100.0%) |  |
| AKT3 Mutation |  |  |  |
| Present | 2 (2.7%) | 3 (5.8%) | 0.6484 |
| Absent | 71 (97.3%) | 49 (94.2%) |  |
| PPP2R1A Mutation |  |  |  |
| Present | 0 (0.0%) | 4 (7.7%) | 0.02793 |
| Absent | 73 (100.0%) | 48 (92.3%) |  |
| TSC1 Mutation |  |  |  |
| Present | 2 (2.7%) | 6 (11.5%) | 0.06586 |
| Absent | 71 (97.3%) | 46 (88.5%) |  |
| TSC2 Mutation |  |  |  |
| Present | 3 (4.1%) | 5 (9.6%) | 0.2753 |
| Absent | 70 (95.9%) | 47 (90.4%) |  |
| STK11 Mutation |  |  |  |
| Present | 0 (0.0%) | 0 (0.0%) | 1 |
| Absent | 73 (100.0%) | 52 (100.0%) |  |
| RHEB Mutation |  |  |  |
| Present | 0 (0.0%) | 0 (0.0%) | 1 |
| Absent | 73 (100.0%) | 52 (100.0%) |  |
| RICTOR Mutation |  |  |  |
| Present | 0 (0.0%) | 2 (3.8%) | 0.1711 |
| Absent | 73 (100.0%) | 50 (96.2%) |  |
| MTOR Mutation |  |  |  |
| Present | 4 (5.5%) | 4 (7.7%) | 0.7178 |
| Absent | 69 (94.5%) | 48 (92.3%) |  |
| RPTOR Mutation |  |  |  |
| Present | 2 (2.7%) | 4 (7.7%) | 0.2328 |
| Absent | 71 (97.3%) | 48 (92.3%) |  |

**Table S2.** LO HL Treated with FOLFOX v LO HL Not Treated with FOLFOX.

| Gene | Late-Onset Hispanic/Latino<br>Treated with FOLFOX<br>n (%) | Late-Onset Hispanic/Latino<br>Not Treated with FOLFOX<br>n (%) | p-value |
| --- | --- | --- | --- |
| PTEN Mutation |  |  |  |
| Present | 4 (4.4%) | 3 (6.0%) | 0.6985 |
| Absent | 87 (95.6%) | 47 (94.0%) |  |
| PIK3R1 Mutation |  |  |  |
| Present | 4 (4.4%) | 0 (0.0%) | 0.297 |
| Absent | 87 (95.6%) | 50 (100.0%) |  |
| PIK3R2 Mutation |  |  |  |
| Present | 1 (1.1%) | 3 (6.0%) | 0.1276 |
| Absent | 90 (98.9%) | 47 (94.0%) |  |
| PIK3R3 Mutation |  |  |  |
| Present | 1 (1.1%) | 1 (2.0%) | 1 |
| Absent | 90 (98.9%) | 49 (98.0%) |  |
| PIK3CA Mutation |  |  |  |
| Present | 24 (26.4%) | 9 (18.0%) | 0.3599 |
| Absent | 67 (73.6%) | 41 (82.0%) |  |
| INPP4B Mutation |  |  |  |
| Present | 1 (1.1%) | 0 (0.0%) | 1 |
| Absent | 90 (98.9%) | 50 (100.0%) |  |
| AKT1 Mutation |  |  |  |
| Present | 2 (2.2%) | 2 (4.0%) | 0.615 |
| Absent | 89 (97.8%) | 48 (96.0%) |  |
| AKT2 Mutation |  |  |  |
| Present | 2 (2.2%) | 0 (0.0%) | 0.539 |
| Absent | 89 (97.8%) | 50 (100.0%) |  |
| AKT3 Mutation |  |  |  |
| Present | 2 (2.2%) | 2 (4.0%) | 0.615 |
| Absent | 89 (97.8%) | 48 (96.0%) |  |
| PPP2R1A Mutation |  |  |  |
| Present | 0 (0.0%) | 2 (4.0%) | 0.1241 |
| Absent | 91 (100.0%) | 48 (96.0%) |  |
| TSC1 Mutation |  |  |  |
| Present | 2 (2.2%) | 2 (4.0%) | 0.615 |
| Absent | 89 (97.8%) | 48 (96.0%) |  |
| TSC2 Mutation |  |  |  |
| Present | 6 (6.6%) | 1 (2.0%) | 0.4213 |
| Absent | 85 (93.4%) | 49 (98.0%) |  |
| STK11 Mutation |  |  |  |
| Present | 0 (0.0%) | 0 (0.0%) | 1 |
| Absent | 91 (100.0%) | 50 (100.0%) |  |
| RHEB Mutation |  |  |  |
| Present | 0 (0.0%) | 0 (0.0%) | 1 |
| Absent | 91 (100.0%) | 50 (100.0%) |  |
| RICTOR Mutation |  |  |  |
| Present | 0 (0.0%) | 4 (8.0%) | 0.0146 |
| Absent | 91 (100.0%) | 46 (92.0%) |  |
| MTOR Mutation |  |  |  |
| Present | 4 (4.4%) | 3 (6.0%) | 0.6985 |
| Absent | 87 (95.6%) | 47 (94.0%) |  |
| RPTOR Mutation |  |  |  |
| Present | 0 (0.0%) | 0 (0.0%) | 1 |
| Absent | 91 (100.0%) | 50 (100.0%) |  |

**Table S3.** EO NHW Treated with FOLFOX v EO NHW Not Treated with FOLFOX.

| Gene | Early-Onset NHW<br>Treated with FOLFOX<br>n (%) | Early-Onset NHW<br>Not Treated with FOLFOX<br>n (%) | p-value |
| --- | --- | --- | --- |
| PTEN Mutation |  |  |  |
| Present | 19 (5.1%) | 23 (7.6%) | 0.2276 |
| Absent | 356 (94.9%) | 279 (92.4%) |  |
| PIK3R1 Mutation |  |  |  |
| Present | 20 (5.3%) | 25 (8.3%) | 0.1695 |
| Absent | 355 (94.7%) | 277 (91.7%) |  |
| PIK3R2 Mutation |  |  |  |
| Present | 6 (1.6%) | 8 (2.6%) | 0.4954 |
| Absent | 369 (98.4%) | 294 (97.4%) |  |
| PIK3R3 Mutation |  |  |  |
| Present | 4 (1.1%) | 4 (1.3%) | 1 |
| Absent | 371 (98.9%) | 298 (98.7%) |  |
| PIK3CA Mutation |  |  |  |
| Present | 72 (19.2%) | 65 (21.5%) | 0.5146 |
| Absent | 303 (80.8%) | 237 (78.5%) |  |
| INPP4B Mutation |  |  |  |
| Present | 5 (1.3%) | 3 (1.0%) | 0.7376 |
| Absent | 370 (98.7%) | 299 (99.0%) |  |
| AKT1 Mutation |  |  |  |
| Present | 9 (2.4%) | 4 (1.3%) | 0.4034 |
| Absent | 366 (97.6%) | 298 (98.7%) |  |
| AKT2 Mutation |  |  |  |
| Present | 2 (0.5%) | 2 (0.7%) | 1 |
| Absent | 373 (99.5%) | 300 (99.3%) |  |
| AKT3 Mutation |  |  |  |
| Present | 3 (0.8%) | 9 (3.0%) | 0.04069 |
| Absent | 372 (99.2%) | 293 (97.0%) |  |
| PPP2R1A Mutation |  |  |  |
| Present | 9 (2.4%) | 9 (3.0%) | 0.8211 |
| Absent | 366 (97.6%) | 293 (97.0%) |  |
| TSC1 Mutation |  |  |  |
| Present | 7 (1.9%) | 10 (3.3%) | 0.3436 |
| Absent | 368 (98.1%) | 292 (96.7%) |  |
| TSC2 Mutation |  |  |  |
| Present | 11 (2.9%) | 14 (4.6%) | 0.3558 |
| Absent | 364 (97.1%) | 288 (95.4%) |  |
| STK11 Mutation |  |  |  |
| Present | 2 (0.5%) | 6 (2.0%) | 0.1484 |
| Absent | 373 (99.5%) | 296 (98.0%) |  |
| RHEB Mutation |  |  |  |
| Present | 3 (0.8%) | 0 (0.0%) | 0.2576 |
| Absent | 372 (99.2%) | 302 (100.0%) |  |
| RICTOR Mutation |  |  |  |
| Present | 7 (1.9%) | 11 (3.6%) | 0.2351 |
| Absent | 368 (98.1%) | 291 (96.4%) |  |
| MTOR Mutation |  |  |  |
| Present | 16 (4.3%) | 19 (6.3%) | 0.3134 |
| Absent | 359 (95.7%) | 283 (93.7%) |  |
| RPTOR Mutation |  |  |  |
| Present | 10 (2.7%) | 6 (2.0%) | 0.7456 |
| Absent | 365 (97.3%) | 296 (98.0%) |  |

**Table S4.** LO NHW Treated with FOLFOX v LO NHW Not Treated with FOLFOX.

| Gene | Late-Onset NHW<br>Treated with FOLFOX<br>n (%) | Late-Onset NHW<br>Not Treated with FOLFOX<br>n (%) | p-value |
| --- | --- | --- | --- |
| PTEN Mutation |  |  |  |
| Present | 45 (4.9%) | 40 (6.1%) | 0.3428 |
| Absent | 874 (95.1%) | 613 (93.9%) |  |
| PIK3R1 Mutation |  |  |  |
| Present | 43 (4.7%) | 37 (5.7%) | 0.4466 |
| Absent | 876 (95.3%) | 616 (94.3%) |  |
| PIK3R2 Mutation |  |  |  |
| Present | 15 (1.6%) | 18 (2.8%) | 0.1758 |
| Absent | 904 (98.4%) | 635 (97.2%) |  |
| PIK3R3 Mutation |  |  |  |
| Present | 5 (0.5%) | 8 (1.2%) | 0.2353 |
| Absent | 914 (99.5%) | 645 (98.8%) |  |
| PIK3CA Mutation |  |  |  |
| Present | 192 (20.9%) | 149 (22.8%) | 0.3949 |
| Absent | 727 (79.1%) | 504 (77.2%) |  |
| INPP4B Mutation |  |  |  |
| Present | 13 (1.4%) | 19 (2.9%) | 0.05912 |
| Absent | 906 (98.6%) | 634 (97.1%) |  |
| AKT1 Mutation |  |  |  |
| Present | 16 (1.7%) | 19 (2.9%) | 0.1694 |
| Absent | 903 (98.3%) | 634 (97.1%) |  |
| AKT2 Mutation |  |  |  |
| Present | 6 (0.7%) | 12 (1.8%) | 0.05296 |
| Absent | 913 (99.3%) | 641 (98.2%) |  |
| AKT3 Mutation |  |  |  |
| Present | 9 (1.0%) | 12 (1.8%) | 0.2158 |
| Absent | 910 (99.0%) | 641 (98.2%) |  |
| PPP2R1A Mutation |  |  |  |
| Present | 17 (1.8%) | 23 (3.5%) | 0.05582 |
| Absent | 902 (98.2%) | 630 (96.5%) |  |
| TSC1 Mutation |  |  |  |
| Present | 17 (1.8%) | 18 (2.8%) | 0.3043 |
| Absent | 902 (98.2%) | 635 (97.2%) |  |
| TSC2 Mutation |  |  |  |
| Present | 30 (3.3%) | 32 (4.9%) | 0.1308 |
| Absent | 889 (96.7%) | 621 (95.1%) |  |
| STK11 Mutation |  |  |  |
| Present | 12 (1.3%) | 15 (2.3%) | 0.1957 |
| Absent | 907 (98.7%) | 638 (97.7%) |  |
| RHEB Mutation |  |  |  |
| Present | 1 (0.1%) | 1 (0.2%) | 1 |
| Absent | 918 (99.9%) | 652 (99.8%) |  |
| RICTOR Mutation |  |  |  |
| Present | 24 (2.6%) | 17 (2.6%) | 1 |
| Absent | 895 (97.4%) | 636 (97.4%) |  |
| MTOR Mutation |  |  |  |
| Present | 41 (4.5%) | 51 (7.8%) | 0.007398 |
| Absent | 878 (95.5%) | 602 (92.2%) |  |
| RPTOR Mutation |  |  |  |
| Present | 19 (2.1%) | 22 (3.4%) | 0.1513 |
| Absent | 900 (97.9%) | 631 (96.6%) |  |

**Table S5.** EO HL Treated with FOLFOX v EO NHW Treated with FOLFOX.

| Gene | Early-Onset Hispanic/Latino<br>Treated with FOLFOX<br>n (%) | Early-Onset NHW<br>Treated with FOLFOX<br>n (%) | p-value |
| --- | --- | --- | --- |
| PTEN Mutation |  |  |  |
| Present | 4 (5.5%) | 19 (5.1%) | 0.7777 |
| Absent | 69 (94.5%) | 356 (94.9%) |  |
| PIK3R1 Mutation |  |  |  |
| Present | 1 (1.4%) | 20 (5.3%) | 0.2238 |
| Absent | 72 (98.6%) | 355 (94.7%) |  |
| PIK3R2 Mutation |  |  |  |
| Present | 5 (6.8%) | 6 (1.6%) | 0.02521 |
| Absent | 68 (93.2%) | 369 (98.4%) |  |
| PIK3R3 Mutation |  |  |  |
| Present | 1 (1.4%) | 4 (1.1%) | 0.5909 |
| Absent | 72 (98.6%) | 371 (98.9%) |  |
| PIK3CA Mutation |  |  |  |
| Present | 14 (19.2%) | 72 (19.2%) | 1 |
| Absent | 59 (80.8%) | 303 (80.8%) |  |
| INPP4B Mutation |  |  |  |
| Present | 0 (0.0%) | 5 (1.3%) | 1 |
| Absent | 73 (100.0%) | 370 (98.7%) |  |
| AKT1 Mutation |  |  |  |
| Present | 4 (5.5%) | 9 (2.4%) | 0.2414 |
| Absent | 69 (94.5%) | 366 (97.6%) |  |
| AKT2 Mutation |  |  |  |
| Present | 0 (0.0%) | 2 (0.5%) | 1 |
| Absent | 73 (100.0%) | 373 (99.5%) |  |
| AKT3 Mutation |  |  |  |
| Present | 2 (2.7%) | 3 (0.8%) | 0.1883 |
| Absent | 71 (97.3%) | 372 (99.2%) |  |
| PPP2R1A Mutation |  |  |  |
| Present | 0 (0.0%) | 9 (2.4%) | 0.3664 |
| Absent | 73 (100.0%) | 366 (97.6%) |  |
| TSC1 Mutation |  |  |  |
| Present | 2 (2.7%) | 7 (1.9%) | 0.6445 |
| Absent | 71 (97.3%) | 368 (98.1%) |  |
| TSC2 Mutation |  |  |  |
| Present | 3 (4.1%) | 11 (2.9%) | 0.7102 |
| Absent | 70 (95.9%) | 364 (97.1%) |  |
| STK11 Mutation |  |  |  |
| Present | 0 (0.0%) | 2 (0.5%) | 1 |
| Absent | 73 (100.0%) | 373 (99.5%) |  |
| RHEB Mutation |  |  |  |
| Present | 0 (0.0%) | 3 (0.8%) | 1 |
| Absent | 73 (100.0%) | 372 (99.2%) |  |
| RICTOR Mutation |  |  |  |
| Present | 0 (0.0%) | 7 (1.9%) | 0.6049 |
| Absent | 73 (100.0%) | 368 (98.1%) |  |
| MTOR Mutation |  |  |  |
| Present | 4 (5.5%) | 16 (4.3%) | 0.5494 |
| Absent | 69 (94.5%) | 359 (95.7%) |  |
| RPTOR Mutation |  |  |  |
| Present | 2 (2.7%) | 10 (2.7%) | 1 |
| Absent | 71 (97.3%) | 365 (97.3%) |  |

**Table S6.** EO HL Not Treated with FOLFOX v EO NHW Not Treated with FOLFOX.

| Gene | Early-Onset Hispanic/Latino<br>Not Treated with FOLFOX<br>n (%) | Early-Onset NHW<br>Not Treated with FOLFOX<br>n (%) | p-value |
| --- | --- | --- | --- |
| PTEN Mutation |  |  |  |
| Present | 8 (15.4%) | 23 (7.6%) | 0.1176 |
| Absent | 44 (84.6%) | 279 (92.4%) |  |
| PIK3R1 Mutation |  |  |  |
| Present | 3 (5.8%) | 25 (8.3%) | 0.4809 |
| Absent | 49 (94.2%) | 277 (91.7%) |  |
| PIK3R2 Mutation |  |  |  |
| Present | 1 (1.9%) | 8 (2.6%) | 1 |
| Absent | 51 (98.1%) | 294 (97.4%) |  |
| PIK3R3 Mutation |  |  |  |
| Present | 1 (1.9%) | 4 (1.3%) | 0.5503 |
| Absent | 51 (98.1%) | 298 (98.7%) |  |
| PIK3CA Mutation |  |  |  |
| Present | 13 (25.0%) | 65 (21.5%) | 0.7057 |
| Absent | 39 (75.0%) | 237 (78.5%) |  |
| INPP4B Mutation |  |  |  |
| Present | 5 (9.6%) | 3 (1.0%) | 0.002262 |
| Absent | 47 (90.4%) | 299 (99.0%) |  |
| AKT1 Mutation |  |  |  |
| Present | 2 (3.8%) | 4 (1.3%) | 0.2153 |
| Absent | 50 (96.2%) | 298 (98.7%) |  |
| AKT2 Mutation |  |  |  |
| Present | 0 (0.0%) | 2 (0.7%) | 1 |
| Absent | 52 (100.0%) | 300 (99.3%) |  |
| AKT3 Mutation |  |  |  |
| Present | 3 (5.8%) | 9 (3.0%) | 0.3954 |
| Absent | 49 (94.2%) | 293 (97.0%) |  |
| PPP2R1A Mutation |  |  |  |
| Present | 4 (7.7%) | 9 (3.0%) | 0.1073 |
| Absent | 48 (92.3%) | 293 (97.0%) |  |
| TSC1 Mutation |  |  |  |
| Present | 6 (11.5%) | 10 (3.3%) | 0.02282 |
| Absent | 46 (88.5%) | 292 (96.7%) |  |
| TSC2 Mutation |  |  |  |
| Present | 5 (9.6%) | 14 (4.6%) | 0.2549 |
| Absent | 47 (90.4%) | 288 (95.4%) |  |
| STK11 Mutation |  |  |  |
| Present | 0 (0.0%) | 6 (2.0%) | 0.598 |
| Absent | 52 (100.0%) | 296 (98.0%) |  |
| RHEB Mutation |  |  |  |
| Present | 0 (0.0%) | 0 (0.0%) | 1 |
| Absent | 52 (100.0%) | 302 (100.0%) |  |
| RICTOR Mutation |  |  |  |
| Present | 2 (3.8%) | 11 (3.6%) | 1 |
| Absent | 50 (96.2%) | 291 (96.4%) |  |
| MTOR Mutation |  |  |  |
| Present | 4 (7.7%) | 19 (6.3%) | 0.7592 |
| Absent | 48 (92.3%) | 283 (93.7%) |  |
| RPTOR Mutation |  |  |  |
| Present | 4 (7.7%) | 6 (2.0%) | 0.04426 |
| Absent | 48 (92.3%) | 296 (98.0%) |  |

**Table S7.**
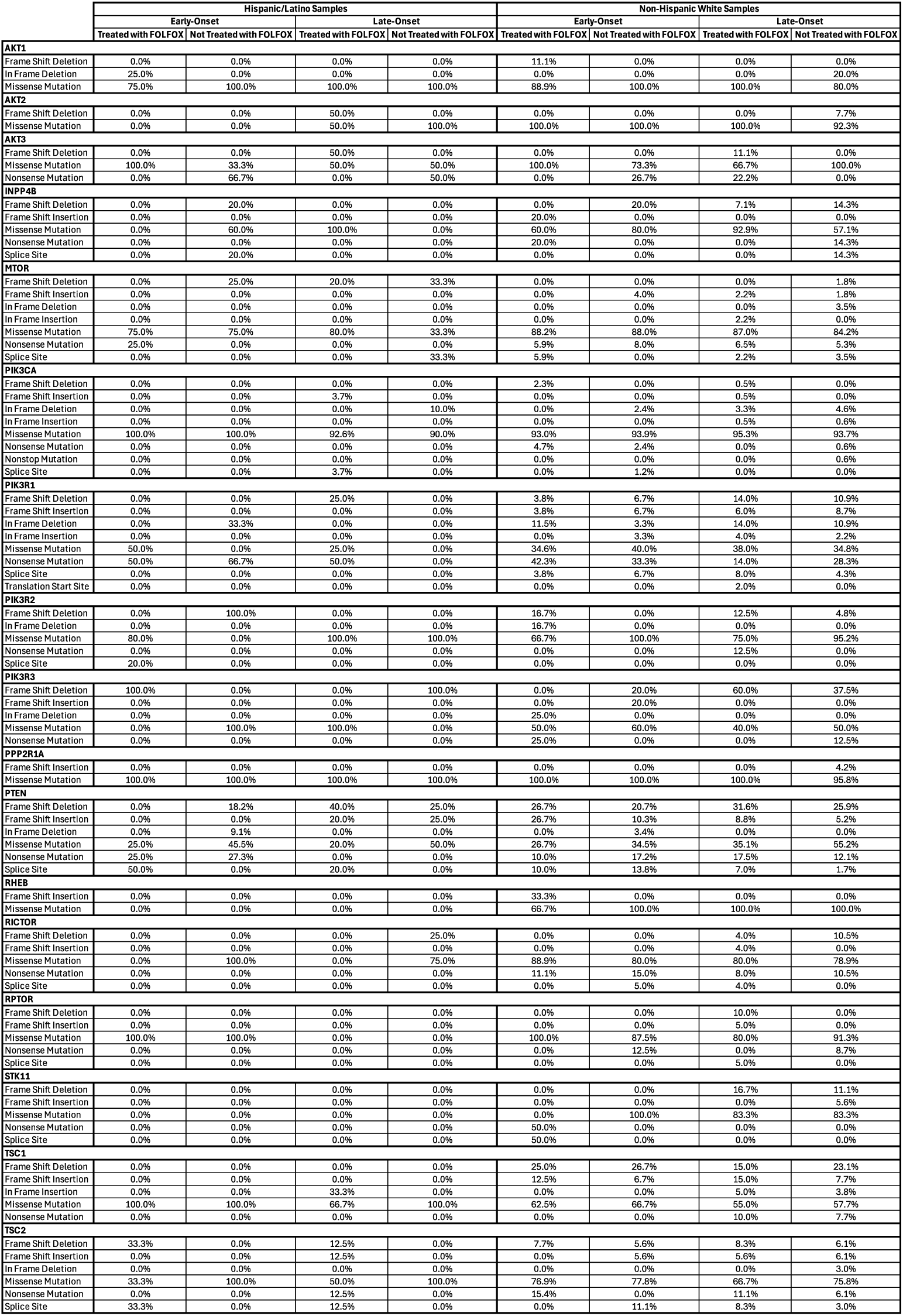
Distribution of TPI3K Pathway Gene Mutation Types by Ancestry, Age of Onset, and FOLFOX Treatment Status in Colorectal Cancer.

**Figure S1.**
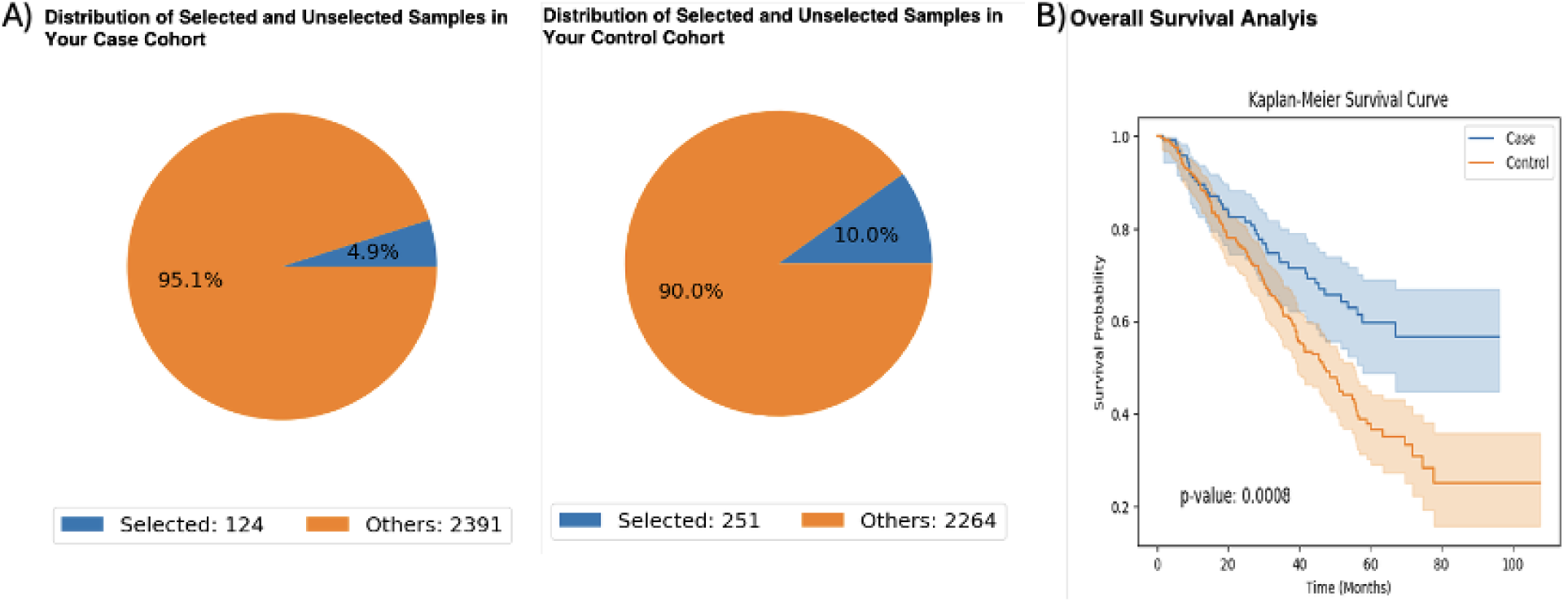
AI-guided selection and survival analysis of early-onset (EO) Non-Hispanic White (NHW) colorectal cancer (CRC) patients treated with FOLFOX, stratified by PI3K pathway alteration status. The AI-HOPE and AI-HOPE-PI3K platforms were used to define case and control cohorts based on integrated clinical, genomic, and treatment criteria. (A) Distribution of selected versus unselected samples in the case cohort—EO NHW CRC patients treated with FOLFOX and harboring PI3K pathway alterations (n = 124)—and the control cohort—EO NHW CRC patients treated with FOLFOX without PI3K pathway alterations (n = 251). (B) Kaplan–Meier overall survival (OS) analysis comparing the two cohorts. Patients with PI3K pathway alterations demonstrated significantly reduced OS compared to those without alterations (log-rank p = 0.0008). Shaded areas represent 95% confidence intervals. Survival curves diverged early, with the altered group showing a steeper decline in OS probability within the first ∼40 months, suggesting a potential negative prognostic impact of PI3K pathway alterations in EO NHW patients receiving FOLFOX chemotherapy.

**Figure S2.**
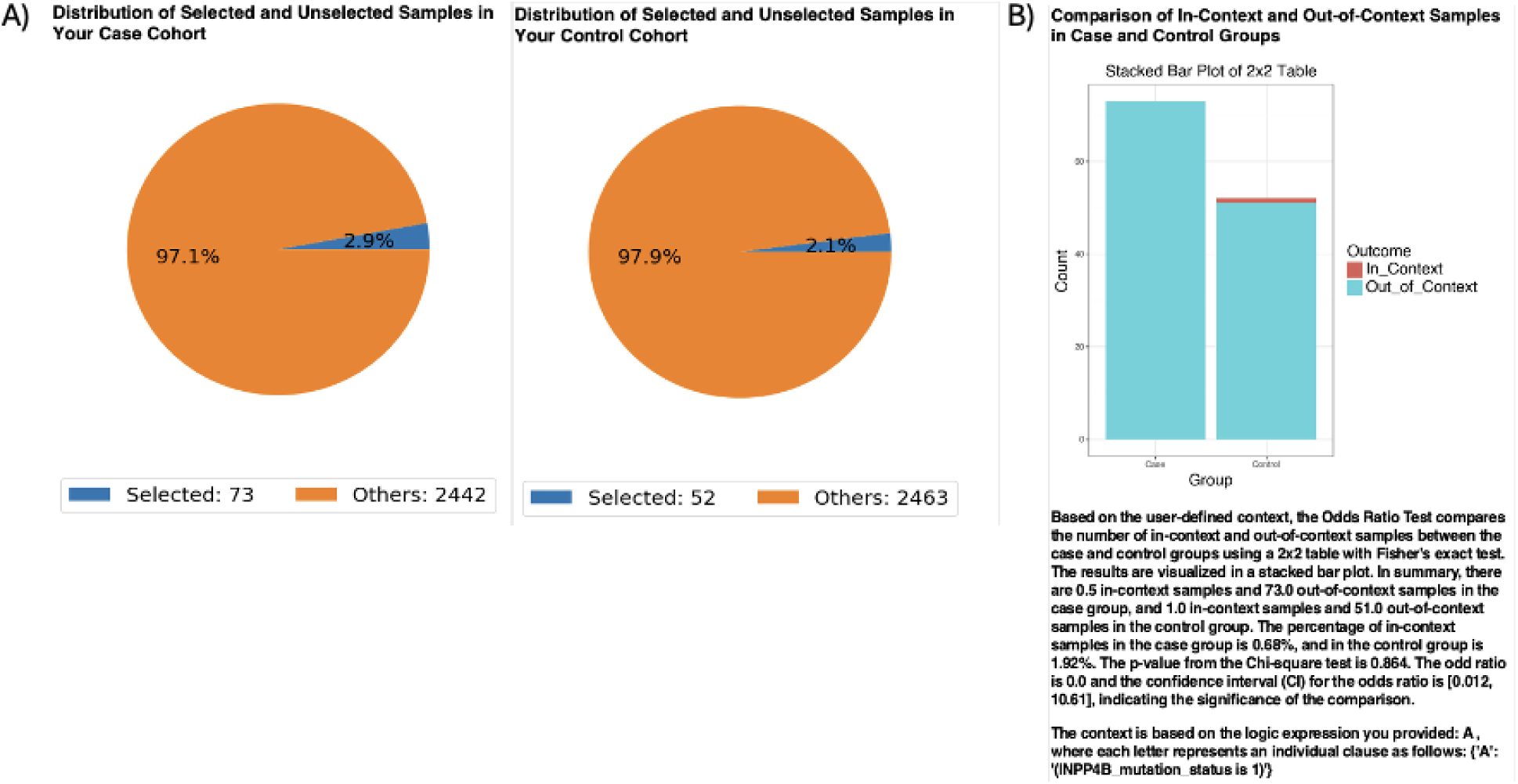
AI-guided selection and odds ratio analysis of early-onset (EO) Hispanic/Latino (H/L) colorectal cancer (CRC) patients with and without FOLFOX treatment, stratified by INPP4B mutation status. The AI-HOPE and AI-HOPE-PI3K platforms were used to define case and control cohorts based on integrated clinical, genomic, and treatment criteria. (A) Distribution of selected versus unselected samples in the case cohort—EO H/L CRC patients treated with FOLFOX (n = 73)—and the control cohort—EO H/L CRC patients not treated with FOLFOX (n = 52). (B) Odds ratio analysis comparing the prevalence of INPP4B mutations between the two cohorts. In-context (mutation-positive) versus out-of-context (mutation-negative) samples were compared using Fisher’s exact test. INPP4B mutations were detected in 6.8% of the case group versus 1.9% of the control group (odds ratio = 3.60; 95% CI: 0.012–10.61; p = 0.864). While not statistically significant, the higher mutation frequency in treated patients suggests a potential association between INPP4B alterations and FOLFOX treatment status in EO H/L CRC.

**Figure S3.**
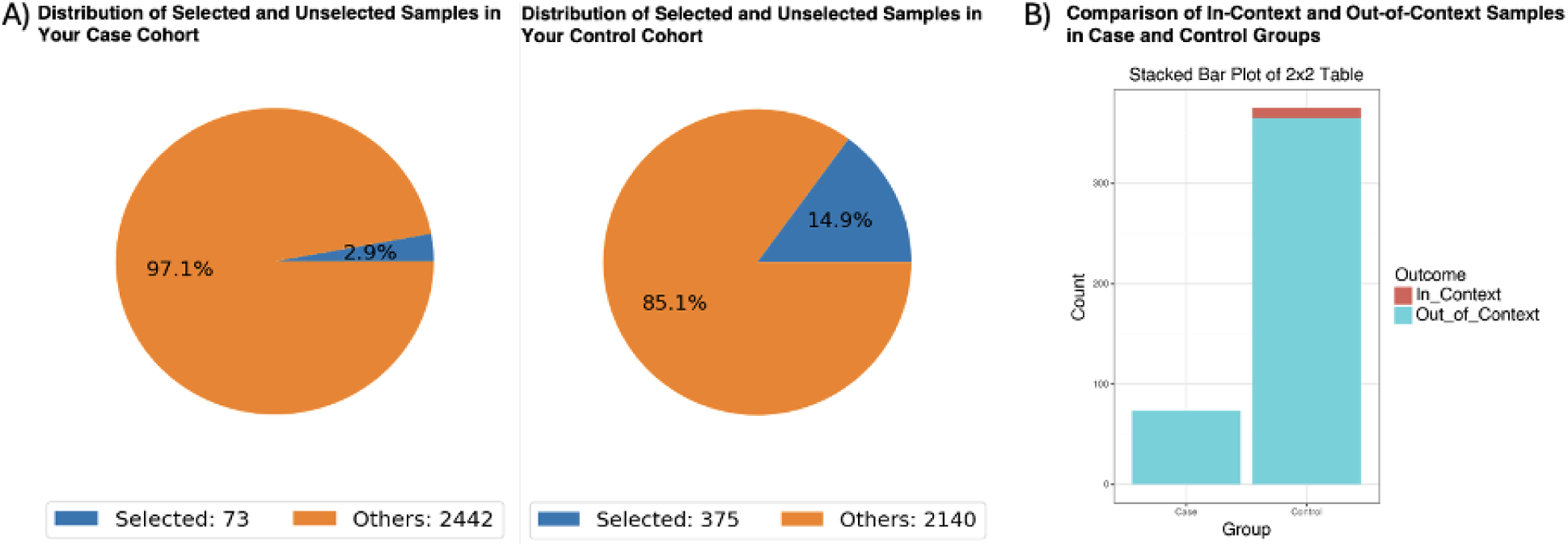
AI-guided selection and mutation prevalence analysis of early-onset (EO) Hispanic/Latino (H/L) versus Non-Hispanic White (NHW) colorectal cancer (CRC) patients not treated with FOLFOX, stratified by RPTOR mutation status. The AI-HOPE and AI-HOPE-PI3K platforms were used to define case and control cohorts based on integrated clinical, genomic, and treatment criteria. The case cohort consisted of EO H/L CRC patients not treated with FOLFOX (n = 73), while the control cohort comprised EO NHW CRC patients not treated with FOLFOX (n = 375). The analysis context was restricted to patients harboring RPTOR mutations. (A) Distribution of selected versus unselected samples in the case and control cohorts, showing the relative representation of patients meeting the inclusion criteria. (B) Odds ratio analysis comparing the prevalence of RPTOR mutations between cohorts. The proportion of mutation-positive cases was 0.68% in the H/L group and 2.67% in the NHW group (odds ratio = 0.00; 95% CI: 0.014–4.328; p = 0.328), indicating no statistically significant difference in mutation prevalence between the groups.

**Figure S4.**
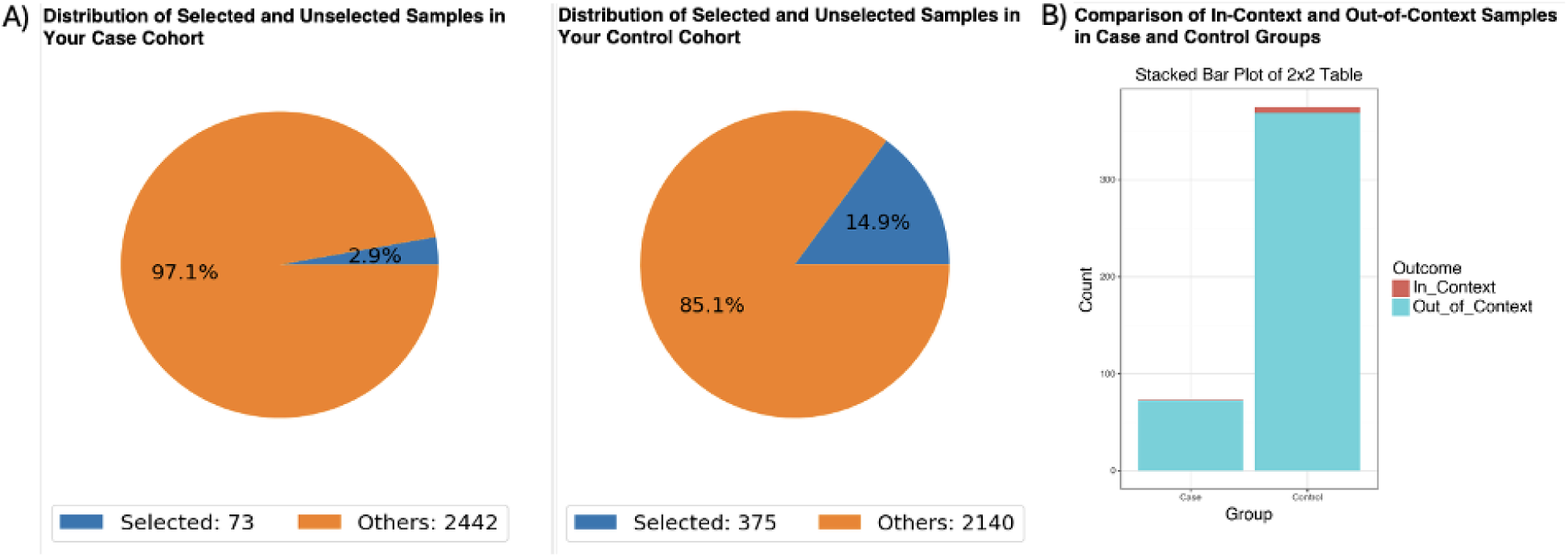
AI-guided cohort selection and comparative analysis of PIK3R2 mutations in early-onset (EO) colorectal cancer (CRC) patients treated with FOLFOX, stratified by ethnicity. The AI-HOPE and AI-HOPE-PI3K platforms were used to define case and control cohorts based on integrated clinical, genomic, and treatment criteria. The case cohort comprised EO Hispanic/Latino (H/L) CRC patients treated with FOLFOX harboring PIK3R2 mutations (n = 73), while the control cohort comprised EO Non-Hispanic White (NHW) CRC patients treated with FOLFOX harboring PIK3R2 mutations (n = 375). (A) Distribution of selected versus unselected samples in the case and control cohorts. The case cohort represented 2.9% of the total dataset (73 of 2,515 CRC patients), while the control cohort represented 14.9% (375 of 2,515 CRC patients).(B) Stacked bar plot of 2×2 table analysis comparing the proportion of in-context samples (mutation present) between cohorts. In-context samples represented 1.37% of the case cohort and 1.6% of the control cohort. Fisher’s exact test yielded an odds ratio of 0.854 (95% CI: 0.101–7.202, p = 1.0), indicating no significant difference in PIK3R2 mutation prevalence between the groups.

**Figure S5.**
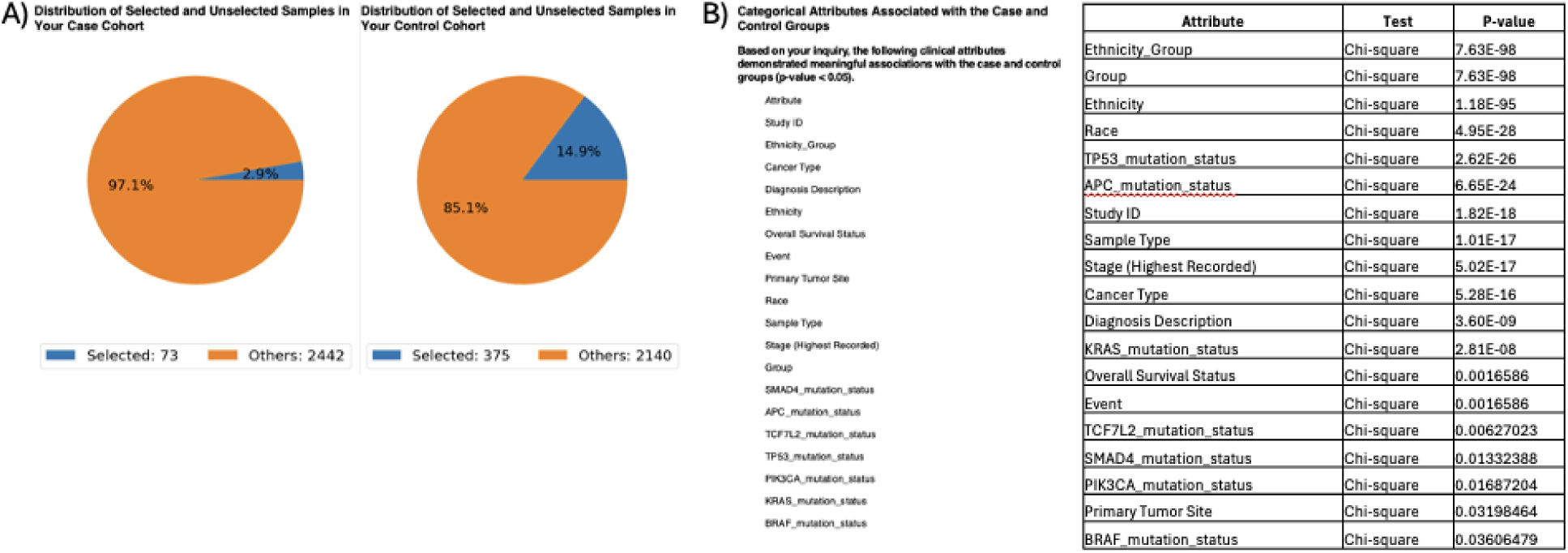
AI-guided cohort selection and identification of significant clinical and genomic differences between early-onset (EO) Hispanic/Latino (H/L) and Non-Hispanic White (NHW) colorectal cancer (CRC) patients treated with FOLFOX. The AI-HOPE and AI-HOPE-PI3K platforms were used to define case and control cohorts based on integrated clinical, genomic, and treatment criteria. The case cohort comprised EO H/L CRC patients treated with FOLFOX (n = 73), while the control cohort comprised EO NHW CRC patients treated with FOLFOX (n = 375). (A) Distribution of selected versus unselected samples in the case and control cohorts, showing the relative proportion of patients meeting the selection criteria compared to the total available dataset. (B) Statistical analysis of categorical clinical and genomic attributes associated with the case and control cohorts. Chi-square testing identified significant differences (p < 0.05) across multiple attributes, including ethnicity, TP53, APC, KRAS, PIK3CA, and BRAF mutation status, tumor stage, cancer type, and overall survival status. The results highlight distinct molecular and clinical profiles between EO H/L and EO NHW CRC patients treated with FOLFOX, underscoring potential ancestry-related differences in tumor biology and treatment response.

